# Adverse childhood experiences as a risk factor for depression-overweight comorbidity in adolescence and young adulthood

**DOI:** 10.1101/2024.07.31.24311274

**Authors:** Fanny Kilpi, Ana L Goncalves Soares, Laura D Howe

**Affiliations:** MRC Integrative Epidemiology Unit at the University of Bristol, UK; Population Health Sciences, Bristol Medical School, University of Bristol, UK

**Keywords:** ALSPAC, depression, body mass index, adolescents

## Abstract

**Background:** The comorbidity of depression and overweight is a manifestation of mental-physical multimorbidity, a marker of complex healthcare needs. We sought to examine how adverse childhood experiences (ACEs) are associated with depression-overweight comorbidity in the period of adolescence and early adulthood, and the extent to which associations are sensitive to age, sex and socioeconomic background.

**Methods:** Using data from 4734 adolescents from the Avon Longitudinal Study of Parents and Children (ALSPAC) birth cohort we estimated relative risk ratios (RRR) for the associations of multiple ACEs (physical, emotional, and sexual abuse, emotional neglect, being bullied, parental substance abuse, violence between parents, parental criminal conviction, parental separation, parental mental illness or suicide) with depression only, overweight only or their comorbidity at ages 17 and 24. We tested whether associations differed by sex and socioeconomic background, indicated by parental education.

**Results:** Most ACEs were associated with depression-overweight comorbidity, and there was a dose-response relationship whereby a greater number of ACEs was associated with greater risk and this continued from adolescence to young adulthood. Some ACEs associations with comorbidity appeared to be influenced by sex: at age 17, females had stronger associations for parental separation and mental health problems, and at age 24, sexual abuse had a stronger association in males. We did not find evidence that the sensitivity to ACEs varied by parental education.

**Conclusions:** ACEs across childhood are associated with depression-overweight comorbidity in late adolescence, which demonstrates their potential impact on the early manifestation of complex healthcare needs.

## Introduction

Adverse childhood experiences (ACEs), such as experiences of neglect and emotional, physical and sexual abuse, are significant risk factors for poor mental and physical health outcomes [1–9]. Evidence from adulthood shows that ACEs are associated with depression [2,4] and higher body mass index (BMI) [7], including early in life [9–13]. Depression and overweight are major contributors to the global burden of ill health [14,15]. They are important public health problems due to the impact that both can have on individual wellbeing as well as societal costs, for example in terms of health spending and economic productivity [16,17]. Depression and overweight tend to emerge in adolescence or young adulthood and carry a high likelihood of tracking into later adulthood [18,19]. Depression and overweight commonly co-occur, which may follow from shared risk factors as well as reciprocal effects between depressive symptoms and BMI [20]. Depression-overweight comorbidity is a manifestation of mental-physical multimorbidity, defined as the presence of at least one mental health condition and at least one physical health condition, that can often emerge already early in the life course. Importantly, mental-physical multimorbidity is a marker of complex healthcare needs, and is strongly linked to socioeconomic circumstances and deprivation [21,22].

ACEs may be connected to the co-occurrence of depression and overweight through the feelings of emotional distress adverse experiences can generate [23]. This can have a direct effect on depressive symptoms and lead to negative coping mechanisms such as emotional overeating [24,25], which could also be a mechanism connecting ACEs to higher BMI. ACEs are linked to poverty and other measures of adverse socioeconomic circumstances [11,26,27], which are associated with both depression and overweight [28]. The differential vulnerability hypothesis posits that socioeconomic disadvantage increases sensitivity to risk exposures such ACEs, as those with fewer socioeconomic resources have fewer of the associated wider resources that could buffer from the negative effects of adversity [29–31].

Previous studies show that socioeconomic position is associated with depression-overweight comorbidity [32], and one previous study found associations between a score of six ACEs and depression-obesity comorbidity at ages 14 and 17 [33]. In this study, we expand on this literature using data from a UK birth cohort to investigate the associations of ACEs with depression-overweight comorbidity in both adolescence and young adulthood with a more comprehensive set of different ACEs. We examine whether the association of ACEs with comorbidity varies across males and females and by family socioeconomic background. We use rich data on ACEs between birth and age 16, reported by the mother and the participants themselves [34], and measures of depression-overweight comorbidity in adolescence (age 17) and early adulthood (age 24).

## Methods

We use data from the Avon Longitudinal Study of Parents and Children (ALSPAC), a population-based birth cohort, which enrolled pregnant women resident in Avon, UK, with expected dates of delivery between 1 April 1991 and 31 December 1992 [35–37]. The initial number of pregnancies enrolled was 14 541, of which 13 988 children were alive at one year. When the oldest children were approximately seven years of age, an attempt was made to bolster the initial sample with eligible cases who had failed to join the study originally. The total sample size for analyses using any data collected after the age of seven is therefore 15 447 pregnancies, of which 14 901 children were alive at one year of age. Study data were collected and managed using REDCap electronic data capture tools hosted at the University of Bristol [38]. REDCap (Research Electronic Data Capture) is a secure, web-based software platform designed to support data capture for research studies. Please note that the study website contains details of all the data that is available through a fully searchable data dictionary and variable search tool (http://www.bristol.ac.uk/alspac/researchers/our-data/). Ethical approval for the study was obtained from the ALSPAC Ethics and Law Committee and the Local Research Ethics Committees. Informed consent for the use of data collected via questionnaires and clinics was obtained from participants following the recommendations of the ALSPAC Ethics and Law Committee at the time.

In 2008-2011, at a target age of 17.5 years, 5217 ALSPAC participants took part in a clinic assessment where weight and height were measured, and 4500 participants responded to an online questionnaire with data on depressive symptoms. A further clinic assessment in 2015-2017 was attended by 4026 participants at approximately 24 years of age, and 4222 ALSPAC participants completed a questionnaire including questions on depressive symptoms at approximately 23 years of age. The selected sample for the present study includes participants who had data on depressive symptoms and/or BMI at the age 17 wave, and at least 10% of ACE questions answered (N=4734 Flow diagram in Supplementary Figure 1).

### Depression-overweight comorbidity

Depression-overweight comorbidity was constructed as a multinomial variable with categories of (i) neither depression nor overweight (reference), (ii) depression only, (iii) overweight only, and (iv) depression-overweight comorbidity. We used the short Moods and Feelings Questionnaire (sMFQ), a 13-item self-reported questionnaire used for screening depressive symptoms in adolescents and young adults [39], to identify depressed mood or depression. Participants were asked to rate statements about experiences of low mood and other correlates in the past two weeks as “not true”, “sometimes” or “true”. Total scores range between zero and 26, with higher scores indicating more depressive symptoms. A score threshold of ≥ 11 was used to identify depression [40]. BMI (kg/m^2^) was calculated from measured weight and height. Height was measured to the nearest millimetre using a Harpenden stadiometer, and weight was measured either to the nearest 0.05kg with a Tanita Body Fat Analyser (age 17) or to the nearest 0.1kg using Tanita TBF-401A electronic body composition scales (age 24). Having overweight was identified with the BMI threshold ≥25kg/m^2^.

### Adverse Childhood Experiences

Several questionnaires throughout the cohort follow up have probed into ACEs. We included ACEs occurring between birth and age 16, and included measures of physical, emotional, and sexual abuse, emotional neglect, being bullied, parental substance abuse, violence between parents, parental conviction of criminal offence, parental separation, parental mental illness or suicide.[34] From age 8 onwards, some ACEs were self-reported by the participants, and at earlier ages, the measures derive mainly from questionnaires filled by the participants’ mothers. The list of the specific questions, who they were reported by and at what ages can be found in a data note by Houtepen and colleagues [34]. We considered the ten different types of ACEs separately with dichotomous indicators of whether the participant reported having experienced the specific type of ACE. We considered any report of an experience to indicate the presence of an ACE, regardless of inconsistencies with later reporting. The number of different types of ACEs was counted and the score was categorized into (i) no ACEs, (ii) one ACE, (iii) two or three ACEs, or (iv) four or more ACEs reported.

### Socioeconomic background

Family socioeconomic background was measured by the highest reported parental educational qualifications from mother and partner responses to questionnaires at 32 weeks’ gestation. We grouped education into (i) university degree, (ii) Advanced (A)-level (examinations around age 18), and (iii) Ordinary (O)-level (examinations taking place at approx. age 16, the UK minimum school leaving age when the mothers were at school), or (iv) CSE (Certificate of Secondary Education), vocational degree or lower qualifications. For interaction analyses, we generated a binary measure of high parental education: (i) high– university degree (ii) low – less than university degree.

### Covariates

Covariates included in adjusted models were ethnicity, social class, financial difficulties, and maternal age (centered at mean age). Ethnicity was reported by the mother (white/non-white). Social class was derived from questionnaires during pregnancy and measured by the highest occupational class based on Registrar General’s Social Class classification: (i) I – professional, (ii) II – managerial/technical, and (ii) III – skilled manual or nonmanual, and (iv) IV – semiskilled manual/V – unskilled manual. Financial difficulties were assessed in a questionnaire at 32 weeks’ gestation, when mothers were asked “How difficult at the moment do you find it to afford these items” for food, heating, and rent or mortgage. We constructed a binary measure of reporting experiencing none or only some difficulties, versus reporting affording any as “fairly difficult” or “very difficult.”

### Missing data

There was missing data on the outcomes, exposures and covariates. After examining patterns of missingness, we decided to impute data with a missing at random (MAR) assumption. Multiple imputation (MI) models included data on all outcomes and variables used in the analysis models, as well as auxiliary variables (Supplementary Table 1), and we stratified the main imputation models by sex and the interaction models by the binary measure of parental education. We generated 50 imputed datasets with 20 cycles and combined coefficients across imputed datasets using Rubin’s rules.

### Statistical methods

We used multinomial logistic regression to examine the associations of the ACE score and different ACEs with depression-overweight comorbidity. We tested for different effects by sex using a joint interaction test using a threshold of P<0.05. To test for interaction between parental education and ACE score, we tested interactions in imputed data that had been stratified by the binary measure of parental education. We also repeated the main analyses in unimputed data as a sensitivity analysis.

## Results

The characteristics of the sample are described in Table 1. A fifth of the participants had no report of any of the ACEs studied, 28% had experienced one ACE, 36% had experienced two or three and 16% had experienced four or more. The most common type of ACE reported was parental mental health problems or suicide (42%), followed by experiences of being bullied (26%) and parental separation (25%). At age 17, 59% of females and 66% of males had neither depression nor overweight, whilst this decreased to 47% for both at age 24 (Table 1). The prevalence of the ‘depression only’ group decreased while the prevalence of ‘overweight only’ and depression-overweight comorbidity increased substantially, the latter from 7% in females and 3% in males to 12% and 8% respectively.

**Table 1.** Characteristics of the unimputed and imputed data (N=4734)

|  | Unimputed<br>All<br>% (N) | All<br>% | Imputed<br>Females<br>% | Males<br>% |
| --- | --- | --- | --- | --- |
| **Sex** |  |  |  |  |
| Male | 44 (2074) | 44 |  |  |
| Female | 56 (2660) | 56 |  |  |
| **Comorbidity at 17** |  |  |  |  |
| Has neither depression nor overweight | 62 (2429) | 62 | 59 | 66 |
| Depression only | 16 (629) | 16 | 19 | 14 |
| Overweight only | 17 (649) | 16 | 16 | 17 |
| Depression-overweight comorbidity | 5 (200) | 5 | 7 | 3 |
| **Comorbidity at 24** |  |  |  |  |
| Has neither depression nor overweight | 51 (1097) | 47 | 47 | 47 |
| Depression only | 13 (278) | 13 | 14 | 11 |
| Overweight only | 27 (579) | 30 | 27 | 34 |
| Depression-overweight comorbidity | 10 (206) | 10 | 12 | 8 |
| **ACEs** |  |  |  |  |
| No ACEs | 22 (620) | 20 | 21 | 20 |
| 1 ACE | 29 (804) | 28 | 26 | 29 |
| 2 to 3 ACEs | 35 (973) | 36 | 37 | 35 |
| 4 or more ACEs | 13 (368) | 16 | 16 | 16 |
| Physical abuse | 17 (635) | 18 | 18 | 17 |
| Sexual abuse | 3 (167) | 4 | 6 | 1 |
| Emotional abuse | 19 (717) | 19 | 20 | 19 |
| Emotional neglect | 18 (716) | 19 | 18 | 20 |
| Being bullied | 26 (1113) | 26 | 23 | 31 |
| Parental substance abuse | 8 (322) | 9 | 9 | 9 |
| Violence between parents | 17 (610) | 19 | 19 | 18 |
| Parental criminal offence | 7 (272) | 7 | 7 | 7 |
| Parental separation | 23 (830) | 25 | 26 | 23 |
| Parental mental health problems or suicide attempt | 41 (1634) | 42 | 43 | 41 |
| **Highest parental education** |  |  |  |  |
| Degree | 30 (1408) | 30 | 29 | 31 |
| A-level | 36 (1668) | 36 | 36 | 36 |
| O-level | 25(1152) | 25 | 24 | 26 |
| CSE / vocational / none | 10 (445) | 10 | 10 | 9 |

The ACE score and several individual ACEs were associated with depression-overweight comorbidity at age 17 in unadjusted results (Supplementary Table 2) as well as when adjusted for socioeconomic covariates (Table 2), with a slight attenuation of associations with adjustment. There was a dose-response relationship between number of ACEs and comorbidity, and participants with four or more ACEs had a nearly threefold increased risk of depression-overweight comorbidity compared to those with no ACEs (RRR 2.92 95% CI 1.71, 5.00) (Table 2).

**Table 2.**
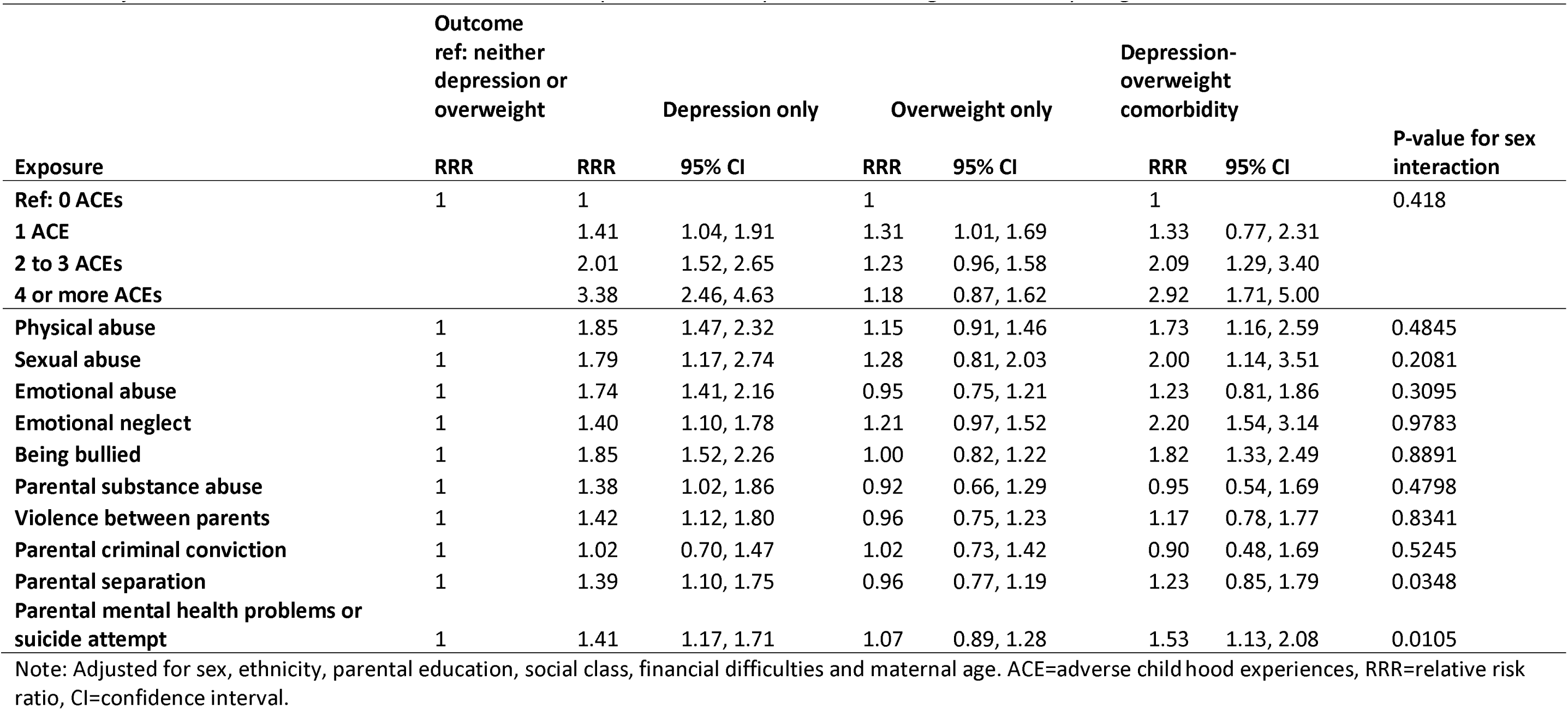
Adjusted associations between adverse childhood experiences and depression-overweight comorbidity at age 17.

When assessing individual ACEs, there was evidence of an association with depression-overweight comorbidity for physical abuse, sexual abuse, emotional neglect, being bullied, and parental mental health problems or suicide attempt (Table 2). The magnitude of the estimates was similar across these ACEs. The associations between ACEs and comorbidity were similar to that of the ‘depression only’ group, while associations with ‘overweight only’ group were weaker or null. For example, the association of physical abuse with depression- overweight comorbidity was RRR 1.73 (95%CI 1.16, 2.59), whilst the association with depression was RRR 1.85 (95%CI 1.47, 2.32) and with overweight RRR 1.15 (95%CI 0.91, 1.46). Tests of interactions by sex suggested that parental separation (P=0.0348) and mental health problems (P=0.0105) had stronger associations with comorbidity in females than males, whilst ACE score and other ACEs did not have different associations by sex (see Supplementary Tables 3 and 4 for sex-specific results).

As with depression-overweight comorbidity at age 17, ACEs were also associated with comorbidity at age 24 in unadjusted models (Supplementary Table 5), with some attenuation of estimates in adjusted models (Table 3). Similarly, comorbidity had more similar estimates to the ‘depression only’ group and ACEs had weaker associations with the ‘overweight only’ group. Four or more ACES were associated with 2.5 times increased risk of comorbidity vs having neither depression or overweight (95% CI 1.58, 3.92) relative to having no ACEs. Of the individual ACEs, physical abuse, sexual abuse, emotional abuse, being bullied, and parental mental health problems were associated with comorbidity at age 24 (Table 3). Sexual abuse was associated with comorbidity particularly strongly for males (P-value for sex difference 0.0094, RRR for female 2.45, 95% CI 1.46,4.11, RRR for males 22.63, 95% CI 3.38,151.74, see Supplementary Tables 6 and 7 for sex-specific results).

**Table 3.** Adjusted associations between adverse childhood experiences and depression-overweight comorbidity at age 24.

|  | **Outcome**<br>**ref: neither**<br>**depression or**<br>**overweight** |  | **Depression only** |  | **Overweight only** |  | **Comorbidity** |  | **P-value for sex**<br>**interaction** |
| --- | --- | --- | --- | --- | --- | --- | --- | --- | --- |
|  | **RRR** | **RRR** | **95% CI** |  | **RRR** | **95% CI** | **RRR** | **95% CI** |  |
| **Ref: 0 ACEs** | 1 | 1 |  |  | 1 |  | 1 |  | 0.9348 |
| **1 ACE** |  | 1.33 | 0.94, 1.88 |  | 1.13 | 0.89, 1.43 | 1.07 | 0.72, 1.58 |  |
| **2 to 3 ACEs** |  | 1.78 | 1.26, 2.51 |  | 1.13 | 0.90, 1.41 | 1.67 | 1.14, 2.45 |  |
| **4 or more ACEs** |  | 2.91 | 1.91, 4.44 |  | 1.07 | 0.80, 1.44 | 2.48 | 1.58, 3.92 |  |
| **Physical abuse** | 1 | 1.95 | 1.44, 2.65 |  | 0.99 | 0.78, 1.25 | 1.93 | 1.44, 2.59 | 0.8871 |
| **Sexual abuse** | 1 | 2.27 | 1.36, 3.78 |  | 1.41 | 0.90, 2.21 | 3.25 | 2.00, 5.28 | 0.0094 |
| **Emotional abuse** | 1 | 1.59 | 1.17, 2.16 |  | 1.03 | 0.82, 1.28 | 1.50 | 1.11, 2.02 | 0.9395 |
| **Emotional neglect** | 1 | 1.04 | 0.77, 1.40 |  | 1.01 | 0.80, 1.26 | 1.29 | 0.94, 1.76 | 0.0707 |
| **Being bullied** | 1 | 1.55 | 1.18, 2.05 |  | 1.00 | 0.83, 1.20 | 1.51 | 1.10, 2.07 | 0.6347 |
| **Parental substance abuse** | 1 | 1.40 | 0.91, 2.15 |  | 0.84 | 0.61, 1.17 | 0.81 | 0.49, 1.34 | 0.7026 |
| **Violence between parents** | 1 | 1.49 | 1.06, 2.09 |  | 1.14 | 0.91, 1.44 | 1.44 | 1.00, 2.06 | 0.993 |
| **Parental criminal conviction** | 1 | 1.30 | 0.83, 2.04 |  | 0.86 | 0.61, 1.22 | 1.28 | 0.76, 2.16 | 0.7463 |
| **Parental separation** | 1 | 1.32 | 0.98, 1.77 |  | 1.00 | 0.81, 1.23 | 1.23 | 0.89, 1.70 | 0.4504 |
| **Parental mental health problems or**<br>**suicide attempt** | 1 | 1.47 | 1.14, 1.90 |  | 1.05 | 0.89, 1.24 | 1.58 | 1.21, 2.08 | 0.1403 |

Note: Adjusted for sex, ethnicity, parental education, social class, financial difficulties and maternal age. ACE=adverse childhood experiences, RRR=relative risk ratio, CI=confidence interval.

The number of ACEs a person had encountered was associated with an increased risk of comorbidity in a dose-response manner in participants with both high and low parental education (Table 4). Stratum-specific estimates were imprecise particularly for high parental education, but there was no statistical evidence of interaction between parental education and ACEs in their association with comorbidity at either age.

**Table 4.** The adjusted risk of having depression-overweigh comorbidity versus having neither depression or overweight associated with ACE score by level of parental education.

|  |  | **Age 17** |  | **Age 24** |  |
| --- | --- | --- | --- | --- | --- |
|  |  | **RRR** | **95% CI** | **RRR** | **95% CI** |
| **Low parental education** | Ref: 0 ACEs | 1 |  | 1 |  |
|  | 1 ACE | 1.29 | 0.66, 2.52 | 1.15 | 0.71, 1.86 |
|  | 2 to 3 ACEs | 2.12 | 1.18, 3.80 | 1.69 | 1.06, 2.70 |
|  | 4 or more ACEs | 3.07 | 1.67, 5.63 | 2.33 | 1.38, 3.95 |
| **High parental education** | Ref: 0 ACEs | 1 |  | 1 |  |
|  | 1 ACE | 1.38 | 0.43, 4.41 | 0.92 | 0.42, 1.99 |
|  | 2 to 3 ACEs | 2.17 | 0.77, 6.11 | 1.46 | 0.72, 2.96 |
|  | 4 or more ACEs | 2.01 | 0.44, 9.10 | 2.40 | 0.97, 5.97 |
| **P-value for interaction with parental education** |  | 0.9329 |  | 0.9904 |  |

Note: Models are adjusted for gender, ethnicity, social class, financial difficulties and maternal age.
ACE=adverse childhood experiences, RRR=relative risk ratio, CI=confidence interval.

The main results using unimputed complete case data were similar to when using the imputed data (Supplementary Table 8, 9 and 10).

## Discussion

We found that ACEs across childhood are associated with depression-overweight comorbidity in adolescence and these associations continue into young adulthood. The risk of depression-overweight comorbidity associated with having four or more ACEs, which comprised 16% of this sample, was over threefold compared to those who reported none. The effects were overall similar at the two ages and in males and females, though a few sex-specific associations were observed. We did not find evidence that parental education moderated the impact of the ACE score on depression-overweight comorbidity.

When exploring the associations between individual ACEs and depression-overweight comorbidity, at age 17, associations were observed for physical abuse, sexual abuse, emotional neglect, being bullied, and parental mental health problems. With the exception of emotional neglect, these associations were also observed at age 24, in addition to associations with emotional abuse and violence between parents. We replicated the associations found between experiences of bullying and parental mental health problems with comorbidity at age 17 found in the Millennium Cohort Study (MCS) (33), although note that we looked at comorbidity of depression and overweight rather than obesity. Having experienced four or more ACEs was a strong risk factor for comorbidity at both ages. The results from MCS had suggested that between ages 14 and 17 the association between ACEs with depression-obesity comorbidity attenuated (33), but used different questionnaires to identify depression at age 14 and 17. We were able to use the same measurement tool for depression at ages 17 and 24 as well as a comprehensive set of ACEs, and the results suggested little attenuation of associations to young adulthood.

There was an increase in comorbidity with age mainly due to an increase in overweight prevalence, especially in males. Nevertheless, the association of ACEs with comorbidity was more similar to the association with depression than the association with overweight status. In our analysis, the associations of the ACE score and the individual ACEs with depression- overweight comorbidity largely mirrored the associations for ‘depression only’, with associations between ACEs and ‘overweight-only’ being small or absent. Overall, we had hypothesized that ACEs may have a strong association with the co-occurrence of depression and overweight, because some of the suggested mechanisms linking stressful experiences with greater BMI, such as impaired self-regulation, poor sleep, overeating and food addiction (23,24), may be heightened by the presence of depression or depressed mood. ACEs could also heighten the relationship between depression and overweight, amplifying their co-occurrence in a ‘vicious cycle’. However, our results indicate that the association with comorbidity at these ages is likely mainly driven by depression. It remains possible that depression could mediate any stronger association between ACEs and overweight emerging later in life, as previous evidence suggests that the impact of ACEs on BMI/obesity is stronger in adulthood than in childhood (5,6). The impact ACEs may have on the intersecting trajectories of depressive symptoms and BMI from adolescence to adulthood would require further investigation.

Despite sex differences in the prevalence of ACEs, depression and overweight, we did not find strong evidence that the association of ACEs with comorbidity varied substantially across sexes. Some differences found for individual ACEs were that parental separation and mental health problems had associations with depression-overweight comorbidity in females but not males at age 17, and sexual abuse had a stronger association with comorbidity in males than females at age 24, although the latter was estimated imprecisely due to small numbers. In the MCS, no sex differences were found for association of cumulative ACE score with depression-obesity comorbidity at age 17 (33). Previous evidence of sex differences in the long-term mental health effects of ACEs has been mixed (2,4,6,7). In the MCS, no differences were found in the association between parent-reported ACEs prior to age 4 and internalizing or externalizing symptoms up to age 14 (13). For overweight status, a systematic review of studies of childhood obesity reported a greater impact of ACEs in females (12).

We also did not find any interaction between parental education and ACEs on their effects on comorbidity, suggesting that resources linked to parental education did not alter sensitivity to ACE exposure or mitigate the negative effects of adversity.

The study strengths include that the data on ACEs throughout childhood have been both prospectively and retrospectively reported. We did not rely solely on parental report of ACEs, which could underestimate their occurrence. Other advantages were that depression was identified based on a validated depressive symptom score rather than a self-reported clinical diagnosis, which could be more strongly affected by socioeconomic biases in seeking care, and overweight was based on measured BMI. However, it should be noted that the measure of depression we used is more sensitive than a clinical diagnosis, and we do not suggest that it corresponds to major depressive disorder as clinically defined. A limitation of the study is the potential bias introduced by study attrition and missing data, which we mitigated using multiple imputation. Nevertheless, the sample remains a more advantaged sample than the general population at the time, which may also result in an underestimation of any ACE effects.

## Conclusions

This study confirms that ACEs are associated with depression-overweight comorbidity in adolescence and young adulthood. The findings highlight the early emergence of complex healthcare needs and mental-physical multimorbidity in a proportion of people exposed to childhood adversity.

## Data Availability

The informed consent obtained from ALSPAC participants does not allow the data to be made freely available through any third party maintained public repository. However, data used for this submission can be made available on request to the ALSPAC Executive. The ALSPAC data management plan describes in detail the policy regarding data sharing, which is through a system of managed open access. Full instructions for applying for data access can be found here: http://www.bristol.ac.uk/alspac/researchers/access/. The ALSPAC study website contains details of all the data that are available (http://www.bristol.ac.uk/alspac/researchers/our-data/).

## Funding

Fanny Kilpi was funded by an Economic and Social Research Council grant (ES/T013923/1). AGS was supported by the Dynamic longitudinal exposome trajectories in cardiovascular and metabolic non-communicable diseases (H2020-SC1-2019-Single-Stage-RTD, project ID 874739). The UK Medical Research Council and Wellcome (Grant ref: 217065/Z/19/Z) and the University of Bristol provide core support for ALSPAC. A comprehensive list of grants funding is available on the ALSPAC website (http://www.bristol.ac.uk/alspac/external/documents/grant-acknowledgements.pdf); The collection of some of the variables used in this study were specifically funded by Wellcome Trust and MRC (076467/Z/05/Z and MR/L022206/1). This publication is the work of the authors and Fanny Kilpi will serve as guarantors for the contents of this paper.

## Conflicts of interest

FK, LDH, AGS: none declared.

## Acknowledgments

We are extremely grateful to all the families who took part in this study, the midwives for their help in recruiting them, and the whole ALSPAC team, which includes interviewers, computer and laboratory technicians, clerical workers, research scientists, volunteers, managers, receptionists and nurses. We thank Dr Lindsey Hines for help with multiple imputation scripts. For the purpose of open access, the author(s) has applied a Creative Commons Attribution (CC BY) licence to any Author Accepted Manuscript version arising from this submission.

## Notes

### Competing Interest Statement

The authors have declared no competing interest.

### Author Declarations

ALSPAC Ethics and Law Committee and the Local Research Ethics Committees gave ethical approval for this work. Informed consent for the use of data was obtained from participants following the recommendations of the ALSPAC Ethics and Law Committee at the time.

