## Supplementary Materials for "Adverse childhood experiences as a risk factor for depression-overweight comorbidity in adolescence and young adulthood"

**Contents**

**Supplementary Figure 1.** Flow diagram of sample selection

**Supplementary Table 1.** Multiple imputation models of outcomes, exposures and covariates

**Supplementary Table 2.** Unadjusted associations between adverse childhood experiences and depression-overweight comorbidity at age 17

**Supplementary Table 3.** Associations between adverse childhood experiences and depression-overweight comorbidity at age 17 in females

**Supplementary Table 4.** Associations between adverse childhood experiences and depression-overweight comorbidity at age 17 in males

**Supplementary Table 5.** Unadjusted associations between adverse childhood experiences and depression-overweight comorbidity at age 24

**Supplementary Table 6.** Associations between adverse childhood experiences and depression-overweight comorbidity at age 24 in females

**Supplementary Table 7.** Associations between adverse childhood experiences and depression-overweight comorbidity at age 24 in males

**Supplementary Table 8.** Associations between adverse childhood experiences and depression-overweight comorbidity at age 17 in complete-case data

**Supplementary Table 9.** Associations between adverse childhood experiences and depression-overweight comorbidity at age 24 in complete-case data

**Supplementary Table 10.** The adjusted risk of having depression-overweigh comorbidity versus having neither depression or overweight associated with ACE score by level of parental education in complete-case data

**Supplementary Figure 1.** Flow diagram of sample selection

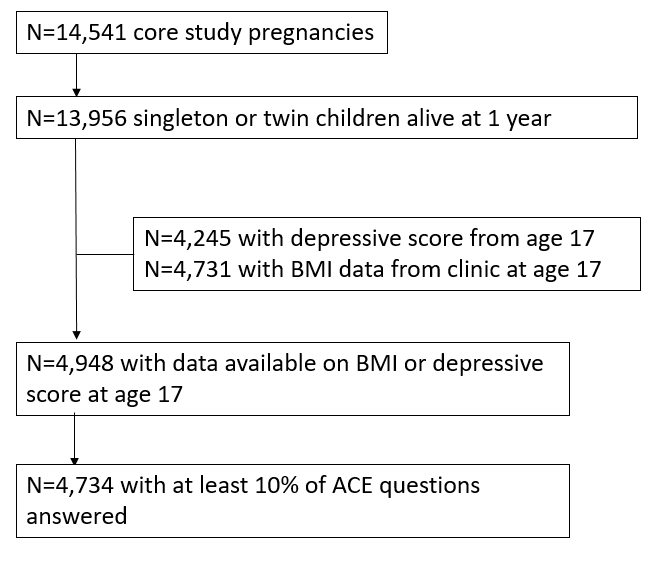

**Supplementary Table 1.** Multiple imputation models of outcomes, exposures and covariates

| **Variable** | **Type** | **Missing data (%)** | **Model** | **Imputation model covariates** |
| --- | --- | --- | --- | --- |
| Sex | covariate | 0 | NA | NA |
| Ethnicity | covariate | 3 | logistic | ACE variables, education, class, financial difficulties, logged BMI at 17, depression at 17, logged BMI at 24, depression at 23, birthweight, gestational age, age of mother, parity, mother's smoking in pregnancy, housing tenure, parents' marital status |
| Physical abuse | exposure | 23 | logistic | ACE variables, social class, financial difficulties, logged BMI at 17, depression at 17, logged BMI at 24, depression at 23, ethnicity, birthweight, gestational age, age of mother, parity, mother's smoking in pregnancy, housing tenure, parents' marital status |
| Sexual abuse | exposure | 6 | logistic | ACE variables, social class, financial difficulties, logged BMI at 17, depression at 17, logged BMI at 24, depression at 23, ethnicity, birthweight, gestational age, age of mother, parity, mother's smoking in pregnancy, housing tenure, parents' marital status |
| Emotional abuse | exposure | 21 | logistic | ACE variables, social class, financial difficulties, logged BMI at 17, depression at 17, logged BMI at 24, depression at 23, ethnicity, birthweight, gestational age, age of mother, parity, mother's smoking in pregnancy, housing tenure, parents' marital status |
| Emotional neglect | exposure | 18 | logistic | ACE variables, social class, financial difficulties, logged BMI at 17, depression at 17, logged BMI at 24, depression at 23, ethnicity, birthweight, gestational age, age of mother, parity, mother's smoking in pregnancy, housing tenure, parents' marital status |
| Being bullied | exposure | 11 | logistic | ACE variables, social class, financial difficulties, logged BMI at 17, depression at 17, logged BMI at 24, depression at 23, ethnicity, birthweight, gestational age, age of mother, parity, mother's smoking in pregnancy, housing tenure, parents' marital status |
| Parental substance abuse | exposure | 19 | logistic | ACE variables, social class, financial difficulties, logged BMI at 17, depression at 17, logged BMI at 24, depression at 23, ethnicity, birthweight, gestational age, age of mother, parity, mother's smoking in pregnancy, housing tenure, parents' marital status |
| Violence between parents | exposure | 26 | logistic | ACE variables, social class, financial difficulties, logged BMI at 17, depression at 17, logged BMI at 24, depression at 23, ethnicity, birthweight, gestational age, age of mother, parity, mother's smoking in pregnancy, housing tenure, parents' marital status |
| Parental criminal offence | exposure | 17 | logistic | ACE variables, social class, financial difficulties, logged BMI at 17, depression at 17, logged BMI at 24, depression at 23, ethnicity, birthweight, gestational age, age of mother, parity, mother's smoking in pregnancy, housing tenure, parents' marital status |
| Parental separation | exposure | 23 | logistic | ACE variables, social class, financial difficulties, logged BMI at 17, depression at 17, logged BMI at 24, depression at 23, ethnicity, birthweight, gestational age, age of mother, parity, mother's smoking in pregnancy, housing tenure, parents' marital status |
| Parental mental health problems or suicide attempt | exposure | 17 | logistic | ACE variables, social class, financial difficulties, logged BMI at 17, depression at 17, logged BMI at 24, depression at 23, ethnicity, birthweight, gestational age, age of mother, parity, mother's smoking in pregnancy, housing tenure, parents' marital status |
| Parental education | covariate | 1 | multinomial logistic | ACE variables, social class, financial difficulties, logged BMI at 17, depression at 17, logged BMI at 24, depression at 23, ethnicity, birthweight, gestational age, age of mother, parity, mother's smoking in pregnancy, housing tenure, parents' marital status |
| Social class | covariate | 2 | multinomial logistic | ACE variables, education, financial difficulties, logged BMI at 17, depression at 17, logged BMI at 24, depression at 23, ethnicity, birthweight, gestational age, age of mother, parity, mother's smoking in pregnancy, housing tenure, parents' marital status |
| Financial difficulties | covariate | 5 | multinomial logistic | education, social class, logged BMI at 17, depression at 17, logged BMI at 24, depression at 23, ethnicity, maternal depressive score, maternal BMI, birthweight, gestational age, age of mother, parity, mother's smoking in pregnancy, housing tenure, parents' marital status |
| Maternal age | covariate | 1 | linear | education, social class, ethnicity |
| Logged BMI at age 17 | outcome | 4 | linear | ACE variables, overweight at age 15, depression at 17, depression at 23, education, class, financial difficulties, ethnicity, birthweight, gestational age, age of mother, parity, mother's smoking in pregnancy, housing tenure, parents' marital status |
| Depression at age 17 | outcome | 13 | logistic | ACE variables, depression at ages 16, 18, 21 and 22, logged BMI at 17, logged BMI at 24, education, class, financial difficulties, ethnicity, birthweight, gestational age, age of mother, parity, mother's smoking in pregnancy, housing tenure, parents' marital status |
| Logged BMI at age 24 | outcome | 39 | linear | ACE variables, overweight at age 15, depression at 17, depression at 23, education, class, financial difficulties, ethnicity, birthweight, gestational age, age of mother, parity, mother's smoking in pregnancy, housing tenure, parents' marital status |
| Depression at age 23 | outcome | 43 | logistic | ACE variables, depression at ages 16, 18, 21 and 22, logged BMI at 17, logged BMI at 24, education, class, financial difficulties, ethnicity, birthweight, gestational age, age of mother, parity, mother's smoking in pregnancy, housing tenure, parents' marital status |

**Supplementary Table 2.** Unadjusted associations between adverse childhood experiences and depression-overweight comorbidity at age 17

|  | **Outcome** |  | |  |  | |  |  | |  |
| --- | --- | --- | --- | --- | --- | --- | --- | --- | --- | --- |
|  | **Ref: neither depression or overweight** | | **Depression only** | | | **Overweight only** | | | **Comorbidity** | |
| **Exposure** | **RRR** | **RRR** | | **95% CI** | **RRR** | | **95% CI** | **RRR** | | **95% CI** |
| **Ref: 0 ACEs** | 1 | 1 | |  | 1 | |  | 1 | |  |
| **1 ACE** |  | 1.44 | | 1.07,1.95 | 1.33 | | 1.03,1.72 | 1.37 | | 0.79,2.37 |
| **2 to 3 ACEs** |  | 2.09 | | 1.59,2.76 | 1.30 | | 1.02,1.67 | 2.34 | | 1.45,3.78 |
| **4 or more ACEs** |  | 3.72 | | 2.74,5.05 | 1.33 | | 0.98,1.81 | 3.65 | | 2.17,6.15 |
| **Physical abuse** | 1 | 1.91 | | 1.53,2.39 | 1.15 | | 0.91,1.46 | 1.76 | | 1.19,2.60 |
| **Sexual abuse** | 1 | 1.95 | | 1.28,2.96 | 1.37 | | 0.87,2.15 | 2.31 | | 1.34,3.98 |
| **Emotional abuse** | 1 | 1.81 | | 1.46,2.24 | 0.96 | | 0.76,1.21 | 1.29 | | 0.86,1.92 |
| **Emotional neglect** | 1 | 1.46 | | 1.16,1.86 | 1.28 | | 1.03,1.59 | 2.53 | | 1.78,3.60 |
| **Being bullied** | 1 | 1.87 | | 1.54,2.27 | 1.01 | | 0.83,1.23 | 1.90 | | 1.39,2.61 |
| **Parental substance abuse** | 1 | 1.51 | | 1.12,2.03 | 1.01 | | 0.73,1.40 | 1.13 | | 0.64,1.98 |
| **Violence between parents** | 1 | 1.51 | | 1.19,1.91 | 1.03 | | 0.81,1.31 | 1.36 | | 0.92,2.01 |
| **Parental criminal conviction** | 1 | 1.06 | | 0.74,1.53 | 1.04 | | 0.75,1.45 | 0.94 | | 0.50,1.75 |
| **Parental separation** | 1 | 1.52 | | 1.22,1.89 | 1.10 | | 0.89,1.36 | 1.58 | | 1.11,2.25 |
| **Parental mental health problems or suicide attempt** | 1 | 1.50 | | 1.25,1.81 | 1.12 | | 0.94,1.34 | 1.69 | | 1.25,2.29 |

Note: Adjusted for sex. ACE=adverse childhood experiences, RRR=relative risk ratio, CI=confidence interval.

**Supplementary Table 3.** Associations between adverse childhood experiences and depression-overweight comorbidity at age 17 in females

|  | **Outcome** |  | |  | |  | |  |  | |  | |  | |  |  | |  | |  | |  |
| --- | --- | --- | --- | --- | --- | --- | --- | --- | --- | --- | --- | --- | --- | --- | --- | --- | --- | --- | --- | --- | --- | --- |
|  | **Ref: Neither depression or overweight** | | **Depression only** | |  | |  | | | **Overweight only** | |  | |  | | | **Comorbidity** | |  | |  | |
|  |  | | **Unadjusted** | | **Adjusted** | |  | | | **Unadjusted** | | **Adjusted** | |  | | | **Unadjusted** | | **Adjusted** | |  | |
| **Exposure** | **RRR** | **RRR** | | **95% CI** | | **RRR** | | **95% CI** | **RRR** | | **95% CI** | | **RRR** | | **95% CI** | **RRR** | | **95% CI** | | **RRR** | | **95% CI** |
| **Ref: 0 ACEs** | 1 | 1 | |  | | 1 | |  | 1 | |  | | 1 | |  | 1 | |  | | 1 | |  |
| **1 ACE** |  | 1.6 | | 1.07,2.41 | | 1.57 | | 1.04,2.37 | 1.74 | | 1.21,2.49 | | 1.69 | | 1.18,2.42 | 1.49 | | 0.78,2.84 | | 1.45 | | 0.76,2.76 |
| **2 to 3 ACEs** |  | 2.45 | | 1.71,3.52 | | 2.36 | | 1.64,3.39 | 1.65 | | 1.17,2.33 | | 1.51 | | 1.07,2.14 | 2.58 | | 1.48,4.49 | | 2.28 | | 1.30,3.99 |
| **4 or more ACEs** |  | 4.78 | | 3.20,7.15 | | 4.29 | | 2.83,6.51 | 1.65 | | 1.07,2.55 | | 1.34 | | 0.86,2.11 | 4.58 | | 2.47,8.50 | | 3.43 | | 1.81,6.49 |
| **Physical abuse** | 1 | 2.13 | | 1.58,2.86 | | 2.05 | | 1.51,2.79 | 1.12 | | 0.80,1.56 | | 1.1 | | 0.79,1.54 | 1.96 | | 1.25,3.07 | | 1.94 | | 1.23,3.06 |
| **Sexual abuse** | 1 | 1.66 | | 1.06,2.60 | | 1.53 | | 0.97,2.42 | 1.23 | | 0.74,2.02 | | 1.12 | | 0.68,1.87 | 1.91 | | 1.05,3.47 | | 1.61 | | 0.87,3.00 |
| **Emotional abuse** | 1 | 2.11 | | 1.62,2.76 | | 2.02 | | 1.54,2.65 | 0.98 | | 0.71,1.36 | | 0.95 | | 0.68,1.31 | 1.36 | | 0.87,2.12 | | 1.26 | | 0.80,2.00 |
| **Emotional neglect** | 1 | 1.46 | | 1.08,1.98 | | 1.37 | | 1.01,1.86 | 1.29 | | 0.95,1.75 | | 1.17 | | 0.86,1.60 | 2.46 | | 1.61,3.75 | | 2.07 | | 1.35,3.17 |
| **Being bullied** | 1 | 1.88 | | 1.45,2.45 | | 1.81 | | 1.38,2.36 | 0.94 | | 0.70,1.27 | | 0.9 | | 0.66,1.22 | 1.97 | | 1.36,2.86 | | 1.78 | | 1.22,2.61 |
| **Parental substance abuse** | 1 | 1.52 | | 1.06,2.19 | | 1.35 | | 0.93,1.96 | 0.81 | | 0.50,1.31 | | 0.7 | | 0.43,1.15 | 0.96 | | 0.50,1.86 | | 0.76 | | 0.39,1.48 |
| **Violence between parents** | 1 | 1.57 | | 1.16,2.12 | | 1.48 | | 1.08,2.01 | 0.97 | | 0.70,1.35 | | 0.87 | | 0.62,1.22 | 1.3 | | 0.82,2.06 | | 1.06 | | 0.65,1.73 |
| **Parental criminal conviction** | 1 | 1.14 | | 0.72,1.81 | | 1.11 | | 0.69,1.77 | 1.22 | | 0.78,1.90 | | 1.21 | | 0.77,1.89 | 1.09 | | 0.54,2.18 | | 1.08 | | 0.53,2.20 |
| **Parental separation** | 1 | 1.67 | | 1.26,2.22 | | 1.54 | | 1.14,2.07 | 1.36 | | 1.02,1.79 | | 1.15 | | 0.86,1.54 | 2.03 | | 1.38,2.99 | | 1.54 | | 1.02,2.34 |
| **Parental mental health problems or suicide attempt** | 1 | 1.79 | | 1.40,2.29 | | 1.69 | | 1.32,2.17 | 1.37 | | 1.07,1.74 | | 1.29 | | 1.01,1.65 | 2.04 | | 1.42,2.92 | | 1.85 | | 1.29,2.67 |

Note: Adjusted for ethnicity, parental education, social class, financial difficulties and maternal age. ACE=adverse childhood experiences, RRR=relative risk ratio, CI=confidence interval.

**Supplementary Table 4.** Associations between adverse childhood experiences and depression-overweight comorbidity at age 17 in males

|  | **Outcome** |  |  |  |  |  |  |  |  |  |  |  |  |
| --- | --- | --- | --- | --- | --- | --- | --- | --- | --- | --- | --- | --- | --- |
|  | **Ref: neither depression or overweight** | **Depression only** | |  |  | **Overweight only** | |  |  | **Comorbidity** | |  |  |
|  |  | **Unadjusted** | | **Adjusted** |  | **Unadjusted** | | **Adjusted** |  | **Unadjusted** | | **Adjusted** |  |
| **Exposure** | **RRR** | **RRR** | **95% CI** | **RRR** | **95% CI** | **RRR** | **95% CI** | **RRR** | **95% CI** | **RRR** | **95% CI** | **RRR** | **95% CI** |
| **Ref: 0 ACEs** | 1 | 1 |  | 1 |  | 1 |  | 1 |  | 1 |  | 1 |  |
| **1 ACE** |  | 1.22 | 0.75,2.00 | 1.19 | 0.73,1.96 | 0.97 | 0.68,1.40 | 0.96 | 0.67,1.38 | 1.14 | 0.37,3.57 | 1.12 | 0.36,3.52 |
| **2 to 3 ACEs** |  | 1.63 | 1.02,2.61 | 1.56 | 0.97,2.49 | 0.99 | 0.69,1.41 | 0.96 | 0.66,1.38 | 1.93 | 0.73,5.08 | 1.81 | 0.68,4.81 |
| **4 or more ACEs** |  | 2.54 | 1.57,4.10 | 2.31 | 1.42,3.77 | 1.05 | 0.67,1.62 | 1.02 | 0.65,1.60 | 2.1 | 0.71,6.24 | 2 | 0.66,6.09 |
| **Physical abuse** | 1 | 1.61 | 1.10,2.34 | 1.56 | 1.06,2.29 | 1.2 | 0.85,1.69 | 1.21 | 0.85,1.72 | 1.27 | 0.53,3.06 | 1.26 | 0.52,3.07 |
| **Sexual abuse** | 1 | 4.94 | 1.63,14.92 | 4.33 | 1.40,13.41 | 2.54 | 0.79,8.22 | 2.62 | 0.80,8.58 | 9.03 | 2.21,36.95 | 8.62 | 2.00,37.10 |
| **Emotional abuse** | 1 | 1.39 | 0.98,1.95 | 1.36 | 0.96,1.93 | 0.93 | 0.65,1.33 | 0.96 | 0.67,1.38 | 1.17 | 0.52,2.61 | 1.22 | 0.54,2.75 |
| **Emotional neglect** | 1 | 1.46 | 1.01,2.13 | 1.43 | 0.98,2.08 | 1.27 | 0.93,1.74 | 1.25 | 0.91,1.73 | 2.73 | 1.43,5.24 | 2.49 | 1.29,4.81 |
| **Being bullied** | 1 | 1.84 | 1.36,2.49 | 1.9 | 1.40,2.59 | 1.06 | 0.81,1.39 | 1.08 | 0.82,1.41 | 1.71 | 0.93,3.14 | 1.81 | 0.97,3.36 |
| **Parental substance abuse** | 1 | 1.44 | 0.88,2.34 | 1.34 | 0.82,2.20 | 1.25 | 0.80,1.94 | 1.21 | 0.77,1.90 | 1.58 | 0.57,4.37 | 1.52 | 0.54,4.27 |
| **Violence between parents** | 1 | 1.4 | 0.97,2.03 | 1.31 | 0.90,1.90 | 1.1 | 0.77,1.56 | 1.06 | 0.74,1.53 | 1.56 | 0.74,3.27 | 1.51 | 0.71,3.23 |
| **Parental criminal conviction** | 1 | 0.96 | 0.53,1.75 | 0.88 | 0.48,1.61 | 0.85 | 0.51,1.44 | 0.83 | 0.49,1.40 | 0.57 | 0.14,2.35 | 0.54 | 0.13,2.28 |
| **Parental separation** | 1 | 1.34 | 0.94,1.90 | 1.21 | 0.84,1.74 | 0.85 | 0.61,1.18 | 0.76 | 0.54,1.07 | 0.69 | 0.30,1.63 | 0.59 | 0.24,1.44 |
| **Parental mental health problems or suicide attempt** | 1 | 1.16 | 0.85,1.57 | 1.08 | 0.79,1.47 | 0.89 | 0.68,1.17 | 0.87 | 0.66,1.15 | 1.08 | 0.58,1.98 | 0.99 | 0.53,1.85 |

Note: Adjusted for ethnicity, parental education, social class, financial difficulties and maternal age. ACE=adverse childhood experiences, RRR=relative risk ratio, CI=confidence interval.

**Supplementary Table 5.** Unadjusted associations between adverse childhood experiences and depression-overweight comorbidity at age 24

|  | **Outcome** |  | |  |  | |  |  | |  |
| --- | --- | --- | --- | --- | --- | --- | --- | --- | --- | --- |
|  | **Ref: neither depression or overweight** | | **Depression only** | | | **Overweight only** | | | **Comorbidity** | |
|  |  | | **Unadjusted** | | | **Unadjusted** | | | **Unadjusted** | |
|  | **RRR** | **RRR** | | **95% CI** | **RRR** | | **95% CI** | **RRR** | | **95% CI** |
| **Ref: 0 ACEs** | 1 | 1 | |  | 1 | |  | 1 | |  |
| **1 ACE** |  | 1.34 | | 0.95,1.90 | 1.15 | | 0.91,1.45 | 1.11 | | 0.75,1.63 |
| **2 to 3 ACEs** |  | 1.81 | | 1.28,2.55 | 1.19 | | 0.96,1.48 | 1.83 | | 1.25,2.67 |
| **4 or more ACEs** |  | 3.05 | | 2.01,4.63 | 1.19 | | 0.89,1.59 | 3.01 | | 1.93,4.71 |
| **Physical abuse** | 1 | 1.97 | | 1.47,2.65 | 0.98 | | 0.78,1.23 | 1.93 | | 1.46,2.56 |
| **Sexual abuse** | 1 | 2.34 | | 1.41,3.88 | 1.47 | | 0.94,2.29 | 3.56 | | 2.25,5.65 |
| **Emotional abuse** | 1 | 1.61 | | 1.19,2.19 | 1.02 | | 0.82,1.26 | 1.52 | | 1.13,2.04 |
| **Emotional neglect** | 1 | 1.07 | | 0.80,1.43 | 1.07 | | 0.86,1.34 | 1.43 | | 1.06,1.94 |
| **Being bullied** | 1 | 1.54 | | 1.17,2.04 | 1.01 | | 0.84,1.21 | 1.56 | | 1.14,2.12 |
| **Parental substance abuse** | 1 | 1.51 | | 1.00,2.29 | 0.93 | | 0.67,1.28 | 0.99 | | 0.61,1.63 |
| **Violence between parents** | 1 | 1.54 | | 1.10,2.16 | 1.21 | | 0.96,1.53 | 1.62 | | 1.14,2.32 |
| **Parental criminal conviction** | 1 | 1.32 | | 0.85,2.06 | 0.88 | | 0.62,1.24 | 1.32 | | 0.79,2.21 |
| **Parental separation** | 1 | 1.43 | | 1.07,1.91 | 1.15 | | 0.94,1.41 | 1.56 | | 1.15,2.12 |
| **Parental mental health problems or suicide attempt** | 1 | 1.51 | | 1.18,1.94 | 1.1 | | 0.93,1.30 | 1.74 | | 1.33,2.28 |

Note: Adjusted for sex. ACE=adverse childhood experiences, RRR=relative risk ratio, CI=confidence interval.

**Supplementary Table 6.** Associations between adverse childhood experiences and depression-overweight comorbidity at age 24 in females

|  | **Outcome** |  | |  | |  | |  | |  | |  | |  | |  | |  | |  |  |  |
| --- | --- | --- | --- | --- | --- | --- | --- | --- | --- | --- | --- | --- | --- | --- | --- | --- | --- | --- | --- | --- | --- | --- |
|  | **Ref: Neither depression or overweight** | | **Depression only** | |  | |  | |  | | **Overweight only** | |  | |  | | **Comorbidity** | |  | |  |  |
|  |  | **Unadjusted** | |  | | **Adjusted** | |  | | **Unadjusted** | |  | | **Adjusted** | |  | | **Unadjusted** | |  | **Adjusted** |  |
|  | **RRR** | **RRR** | | **95% CI** | | **RRR** | | **95% CI** | | **RRR** | | **95% CI** | | **RRR** | | **95% CI** | | **RRR** | | **95% CI** | **RRR** | **95% CI** |
| **Ref: 0 ACEs** | 1 | 1 | |  | | 1 | |  | | 1 | |  | | 1 | |  | | 1 | |  | 1 |  |
| **1 ACE** |  | 1.46 | | 0.96,2.23 | | 1.46 | | 0.95,2.22 | | 1.31 | | 0.95,1.80 | | 1.28 | | 0.92,1.77 | | 1.15 | | 0.71,1.85 | 1.11 | 0.69,1.80 |
| **2 to 3 ACEs** |  | 1.92 | | 1.26,2.91 | | 1.92 | | 1.26,2.92 | | 1.43 | | 1.05,1.95 | | 1.31 | | 0.96,1.80 | | 2.05 | | 1.34,3.15 | 1.85 | 1.20,2.86 |
| **4 or more ACEs** |  | 3.18 | | 1.93,5.24 | | 3.10 | | 1.87,5.16 | | 1.31 | | 0.90,1.92 | | 1.06 | | 0.71,1.57 | | 3.22 | | 1.92,5.40 | 2.46 | 1.42,4.28 |
| **Physical abuse** | 1 | 2.02 | | 1.43,2.85 | | 2.01 | | 1.41,2.86 | | 0.91 | | 0.66,1.26 | | 0.89 | | 0.64,1.24 | | 1.81 | | 1.28,2.55 | 1.79 | 1.26,2.54 |
| **Sexual abuse** | 1 | 1.7 | | 0.99,2.91 | | 1.63 | | 0.95,2.81 | | 1.54 | | 0.98,2.42 | | 1.44 | | 0.91,2.29 | | 2.75 | | 1.67,4.51 | 2.45 | 1.46,4.11 |
| **Emotional abuse** | 1 | 1.69 | | 1.17,2.43 | | 1.66 | | 1.15,2.40 | | 1.06 | | 0.79,1.41 | | 1.02 | | 0.76,1.37 | | 1.48 | | 1.03,2.12 | 1.41 | 0.97,2.04 |
| **Emotional neglect** | 1 | 1.37 | | 0.96,1.95 | | 1.32 | | 0.92,1.91 | | 1.09 | | 0.81,1.48 | | 0.97 | | 0.71,1.32 | | 1.87 | | 1.31,2.67 | 1.61 | 1.11,2.32 |
| **Being bullied** | 1 | 1.40 | | 0.99,1.98 | | 1.37 | | 0.97,1.94 | | 1.00 | | 0.76,1.32 | | 0.94 | | 0.71,1.25 | | 1.44 | | 0.99,2.09 | 1.31 | 0.89,1.93 |
| **Parental substance abuse** | 1 | 1.64 | | 1.01,2.67 | | 1.57 | | 0.95,2.59 | | 0.79 | | 0.51,1.22 | | 0.68 | | 0.43,1.06 | | 0.98 | | 0.57,1.68 | 0.77 | 0.44,1.34 |
| **Violence between parents** | 1 | 1.53 | | 1.04,2.25 | | 1.52 | | 1.03,2.25 | | 1.18 | | 0.88,1.58 | | 1.08 | | 0.80,1.45 | | 1.57 | | 1.02,2.40 | 1.35 | 0.86,2.13 |
| **Parental criminal conviction** | 1 | 1.11 | | 0.63,1.96 | | 1.12 | | 0.63,1.98 | | 0.89 | | 0.56,1.42 | | 0.90 | | 0.56,1.43 | | 1.31 | | 0.73,2.35 | 1.33 | 0.73,2.41 |
| **Parental separation** | 1 | 1.23 | | 0.87,1.75 | | 1.16 | | 0.81,1.67 | | 1.21 | | 0.92,1.59 | | 1.01 | | 0.76,1.33 | | 1.49 | | 1.04,2.15 | 1.14 | 0.77,1.67 |
| **Parental mental health problems or suicide attempt** | 1 | 1.66 | | 1.25,2.22 | | 1.64 | | 1.23,2.20 | | 1.29 | | 1.02,1.62 | | 1.22 | | 0.96,1.55 | | 2.11 | | 1.52,2.93 | 1.93 | 1.38,2.69 |

Note: Adjusted for ethnicity, parental education, social class, financial difficulties and maternal age. ACE=adverse childhood experiences, RRR=relative risk ratio, CI=confidence interval.

**Supplementary Table 7.** Associations between adverse childhood experiences and depression-overweight comorbidity at age 24 in males

|  | **Outcome** |  | |  | |  |  |  | |  | |  |  |  | |  | |  |  |
| --- | --- | --- | --- | --- | --- | --- | --- | --- | --- | --- | --- | --- | --- | --- | --- | --- | --- | --- | --- |
|  | **Ref: neither depression or overweight** | | **Depression only** | |  | |  | | **Overweight only** | |  | |  | | **Comorbidity** | |  | |  |
|  |  | | **Unadjusted** | | **Adjusted** | |  | | **Unadjusted** | | **Adjusted** | |  | | **Unadjusted** | | **Adjusted** | |  |
|  | **RRR** | **RRR** | | **95% CI** | | **RRR** | **95% CI** | **RRR** | | **95% CI** | | **RRR** | **95% CI** | **RRR** | | **95% CI** | | **RRR** | **95% CI** |
| **Ref: 0 ACEs** | 1 | 1 | |  | | 1 |  | 1 | |  | | 1 |  | 1 | |  | | 1 |  |
| **1 ACE** |  | 1.18 | | 0.64,2.17 | | 1.16 | 0.63,2.13 | 0.99 | | 0.71,1.37 | | 0.98 | 0.70,1.36 | 1.05 | | 0.52,2.12 | | 1.02 | 0.51,2.07 |
| **2 to 3 ACEs** |  | 1.67 | | 0.94,2.96 | | 1.61 | 0.90,2.88 | 0.96 | | 0.70,1.32 | | 0.94 | 0.68,1.30 | 1.52 | | 0.74,3.14 | | 1.43 | 0.68,2.98 |
| **4 or more ACEs** |  | 2.86 | | 1.40,5.82 | | 2.67 | 1.29,5.51 | 1.06 | | 0.69,1.65 | | 1.05 | 0.67,1.63 | 2.71 | | 1.19,6.17 | | 2.51 | 1.08,5.85 |
| **Physical abuse** | 1 | 1.88 | | 1.11,3.18 | | 1.85 | 1.08,3.17 | 1.06 | | 0.74,1.50 | | 1.09 | 0.76,1.57 | 2.15 | | 1.26,3.67 | | 2.21 | 1.26,3.87 |
| **Sexual abuse** | 1 | 15.31 | | 2.33,100.65 | | 16.51 | 2.39,114.31 | 1.22 | | 0.13,11.98 | | 1.23 | 0.12,12.17 | 21.27 | | 3.35,135.04 | | 22.63 | 3.38,151.74 |
| **Emotional abuse** | 1 | 1.49 | | 0.90,2.46 | | 1.46 | 0.88,2.44 | 0.98 | | 0.70,1.37 | | 1.03 | 0.73,1.45 | 1.60 | | 0.92,2.79 | | 1.68 | 0.94,2.99 |
| **Emotional neglect** | 1 | 0.75 | | 0.44,1.27 | | 0.72 | 0.42,1.25 | 1.03 | | 0.76,1.41 | | 1.02 | 0.75,1.41 | 0.89 | | 0.52,1.53 | | 0.86 | 0.50,1.47 |
| **Being bullied** | 1 | 1.75 | | 1.16,2.63 | | 1.86 | 1.22,2.82 | 1.02 | | 0.79,1.31 | | 1.02 | 0.79,1.32 | 1.74 | | 1.03,2.95 | | 1.82 | 1.06,3.10 |
| **Parental substance abuse** | 1 | 1.28 | | 0.57,2.84 | | 1.13 | 0.49,2.59 | 1.07 | | 0.67,1.72 | | 1.02 | 0.62,1.66 | 0.97 | | 0.36,2.61 | | 0.83 | 0.30,2.28 |
| **Violence between parents** | 1 | 1.55 | | 0.86,2.79 | | 1.42 | 0.78,2.59 | 1.25 | | 0.89,1.76 | | 1.21 | 0.86,1.71 | 1.71 | | 0.91,3.23 | | 1.55 | 0.81,2.97 |
| **Parental criminal conviction** | 1 | 1.68 | | 0.82,3.43 | | 1.61 | 0.78,3.34 | 0.87 | | 0.53,1.43 | | 0.85 | 0.51,1.40 | 1.28 | | 0.49,3.34 | | 1.20 | 0.45,3.20 |
| **Parental separation** | 1 | 1.80 | | 1.09,2.97 | | 1.61 | 0.97,2.67 | 1.08 | | 0.79,1.48 | | 0.98 | 0.71,1.36 | 1.69 | | 0.98,2.93 | | 1.39 | 0.79,2.45 |
| **Parental mental health problems or suicide attempt** | 1 | 1.32 | | 0.85,2.06 | | 1.27 | 0.81,1.99 | 0.92 | | 0.71,1.18 | | 0.90 | 0.70,1.16 | 1.26 | | 0.77,2.07 | | 1.17 | 0.71,1.95 |

Note: Adjusted for ethnicity, parental education, social class, financial difficulties and maternal age. ACE=adverse childhood experiences, RRR=relative risk ratio, CI=confidence interval.

**Supplementary Table 8.** Associations between adverse childhood experiences and depression-overweight comorbidity at age 17 in complete-case data

|  | **Outcome** |  |  |  |  |  |  |  |  |  |  |  |  |  |
| --- | --- | --- | --- | --- | --- | --- | --- | --- | --- | --- | --- | --- | --- | --- |
|  | **Ref: neither depression or overweight** | **Depression only** | |  |  | **Overweight only** | |  |  | **Comorbidity** | |  |  |  |
|  |  | **Unadjusted** | | **Adjusted** |  | **Unadjusted** | | **Adjusted** |  | **Unadjusted** | | **Adjusted** |  |  |
| **Exposure** | **RRR** | **RRR** | **95% CI** | **RRR** | **95% CI** | **RRR** | **95% CI** | **RRR** | **95% CI** | **RRR** | **95% CI** | **RRR** | **95% CI** | **P-value for sex interaction** |
| **Ref: 0 ACEs** | 1 | 1 |  | 1 |  | 1 |  | 1 |  | 1 |  | 1 |  | 0.4104 |
| **1 ACE** |  | 1.25 | 0.86,1.81 | 1.21 | 0.83,1.78 | 1.16 | 0.85,1.58 | 1.13 | 0.82,1.55 | 0.94 | 0.47,1.88 | 0.84 | 0.41,1.71 |  |
| **2 to 3 ACEs** |  | 1.91 | 1.35,2.69 | 1.87 | 1.31,2.67 | 1.14 | 0.84,1.54 | 1.12 | 0.82,1.53 | 2.03 | 1.12,3.67 | 1.77 | 0.97,3.23 |  |
| **4 or more ACEs** |  | 4.42 | 2.99,6.53 | 4.05 | 2.69,6.09 | 1.50 | 1.00,2.23 | 1.45 | 0.96,2.20 | 4.18 | 2.14,8.16 | 3.5 | 1.75,7.00 |  |
| **Physical abuse** | 1 | 2.07 | 1.63,2.63 | 1.98 | 1.54,2.53 | 1.22 | 0.94,1.59 | 1.16 | 0.88,1.52 | 2.05 | 1.37,3.05 | 1.83 | 1.20,2.81 | 0.3812 |
| **Sexual abuse** | 1 | 2.00 | 1.31,3.04 | 1.93 | 1.24,3.00 | 1.41 | 0.88,2.28 | 1.36 | 0.82,2.26 | 2.28 | 1.27,4.10 | 1.83 | 0.95,3.54 | 0.0774 |
| **Emotional abuse** | 1 | 1.87 | 1.49,2.35 | 1.75 | 1.38,2.22 | 0.99 | 0.76,1.28 | 0.96 | 0.74,1.26 | 1.32 | 0.87,1.98 | 1.27 | 0.83,1.94 | 0.2779 |
| **Emotional neglect** | 1 | 1.41 | 1.11,1.79 | 1.39 | 1.09,1.79 | 1.20 | 0.94,1.52 | 1.06 | 0.82,1.38 | 2.46 | 1.73,3.52 | 2.07 | 1.42,3.01 | 0.8198 |
| **Being bullied** | 1 | 1.90 | 1.55,2.32 | 1.83 | 1.48,2.25 | 1.10 | 0.89,1.36 | 1.13 | 0.91,1.40 | 2.01 | 1.44,2.81 | 1.88 | 1.33,2.67 | 0.6684 |
| **Parental substance abuse** | 1 | 1.51 | 1.10,2.08 | 1.42 | 1.01,1.98 | 1.01 | 0.70,1.44 | 1.00 | 0.69,1.44 | 1.22 | 0.69,2.17 | 1.05 | 0.58,1.91 | 0.2478 |
| **Violence between parents** | 1 | 1.52 | 1.18,1.95 | 1.49 | 1.15,1.94 | 0.98 | 0.75,1.28 | 0.97 | 0.73,1.28 | 1.46 | 0.95,2.25 | 1.37 | 0.87,2.15 | 0.6332 |
| **Parental criminal conviction** | 1 | 1.22 | 0.84,1.76 | 1.23 | 0.84,1.79 | 1.18 | 0.82,1.71 | 1.25 | 0.86,1.82 | 1.09 | 0.57,2.06 | 1.02 | 0.52,2.02 | 0.8129 |
| **Parental separation** | 1 | 1.54 | 1.23,1.92 | 1.56 | 1.23,1.97 | 1.08 | 0.86,1.37 | 1.01 | 0.79,1.30 | 1.57 | 1.08,2.29 | 1.21 | 0.81,1.81 | 0.1083 |
| **Parental mental health problems or suicide attempt** | 1 | 1.50 | 1.24,1.82 | 1.37 | 1.12,1.67 | 1.10 | 0.90,1.33 | 1.04 | 0.86,1.27 | 1.61 | 1.16,2.23 | 1.48 | 1.06,2.09 | 0.0096 |

Note: Adjusted for sex, ethnicity, parental education, social class, financial difficulties and maternal age. ACE=adverse childhood experiences, RRR=relative risk ratio, CI=confidence interval.

**Supplementary Table 9.** Associations between adverse childhood experiences and depression-overweight comorbidity at age 24 in complete-case data

|  | **Outcome** |  |  |  |  |  |  |  |  |  |  |  |  |  |
| --- | --- | --- | --- | --- | --- | --- | --- | --- | --- | --- | --- | --- | --- | --- |
|  | **Ref: neither depression or overweight** | **Depression only** | |  |  | **Overweight only** | |  |  | **Comorbidity** | |  |  |  |
|  |  | **Unadjusted** | | **Adjusted** |  | **Unadjusted** | | **Adjusted** |  | **Unadjusted** | | **Adjusted** |  |  |
|  | **RRR** | **RRR** | **95% CI** | **RRR** | **95% CI** | **RRR** | **95% CI** | **RRR** | **95% CI** | **RRR** | **95% CI** | **RRR** | **95% CI** | **P-value for sex interaction** |
| **Ref: 0 ACEs** | 1 | 1 |  | 1 |  | 1 |  | 1 |  | 1 |  | 1 |  | 0.4401 |
| **1 ACE** |  | 1.57 | 0.97,2.54 | 1.63 | 0.98,2.70 | 1.18 | 0.85,1.63 | 1.13 | 0.81,1.59 | 0.69 | 0.38,1.26 | 0.6 | 0.32,1.11 |  |
| **2 to 3 ACEs** |  | 1.92 | 1.21,3.04 | 1.98 | 1.22,3.23 | 1.37 | 1.00,1.87 | 1.37 | 0.99,1.89 | 1.67 | 1.02,2.73 | 1.58 | 0.95,2.62 |  |
| **4 or more ACEs** |  | 3.21 | 1.86,5.56 | 3.32 | 1.86,5.91 | 1.28 | 0.82,1.98 | 1.23 | 0.78,1.95 | 3.09 | 1.74,5.51 | 2.76 | 1.52,5.03 |  |
| **Physical abuse** | 1 | 2.02 | 1.46,2.78 | 2.02 | 1.45,2.83 | 1.12 | 0.85,1.48 | 1.07 | 0.80,1.42 | 2.46 | 1.73,3.50 | 2.20 | 1.52,3.19 | 0.0861 |
| **Sexual abuse** | 1 | 2.12 | 1.21,3.73 | 2.06 | 1.13,3.76 | 1.71 | 1.04,2.82 | 1.71 | 1.01,2.87 | 4.24 | 2.53,7.12 | 4.00 | 2.30,6.95 | 0.0835 |
| **Emotional abuse** | 1 | 1.74 | 1.25,2.43 | 1.74 | 1.23,2.46 | 1.07 | 0.81,1.42 | 1.04 | 0.78,1.39 | 1.77 | 1.22,2.57 | 1.70 | 1.15,2.51 | 0.1021 |
| **Emotional neglect** | 1 | 0.93 | 0.64,1.36 | 0.98 | 0.66,1.45 | 0.98 | 0.74,1.29 | 0.94 | 0.70,1.26 | 1.58 | 1.09,2.29 | 1.50 | 1.02,2.21 | 0.3835 |
| **Being bullied** | 1 | 1.46 | 1.08,1.98 | 1.47 | 1.07,2.03 | 0.91 | 0.71,1.17 | 0.95 | 0.73,1.23 | 1.53 | 1.08,2.16 | 1.40 | 0.97,2.01 | 0.5909 |
| **Parental substance abuse** | 1 | 1.80 | 1.10,2.94 | 1.90 | 1.14,3.17 | 1.15 | 0.75,1.76 | 1.10 | 0.70,1.73 | 0.82 | 0.40,1.68 | 0.72 | 0.34,1.52 | 0.1697 |
| **Violence between parents** | 1 | 1.62 | 1.12,2.36 | 1.63 | 1.10,2.41 | 1.33 | 0.99,1.80 | 1.29 | 0.94,1.77 | 1.81 | 1.20,2.72 | 1.73 | 1.13,2.65 | 0.8114 |
| **Parental criminal conviction** | 1 | 1.23 | 0.73,2.08 | 1.24 | 0.72,2.15 | 0.75 | 0.47,1.18 | 0.82 | 0.52,1.31 | 1.17 | 0.64,2.13 | 1.18 | 0.63,2.23 | 0.511 |
| **Parental separation** | 1 | 1.24 | 0.87,1.75 | 1.19 | 0.83,1.72 | 1.12 | 0.85,1.47 | 0.97 | 0.73,1.29 | 1.29 | 0.87,1.91 | 0.99 | 0.65,1.51 | 0.1683 |
| **Parental mental health problems or suicide attempt** | 1 | 1.49 | 1.12,1.97 | 1.43 | 1.06,1.93 | 1.15 | 0.92,1.44 | 1.13 | 0.90,1.42 | 1.96 | 1.42,2.71 | 1.90 | 1.36,2.66 | 0.1231 |

Note: Adjusted for sex, ethnicity, parental education, social class, financial difficulties and maternal age. ACE=adverse childhood experiences, RRR=relative risk ratio, CI=confidence interval.

**Supplementary Table 10.** The adjusted risk of having depression-overweigh comorbidity versus having neither depression or overweight associated with ACE score by level of parental education in complete-case data

|  |  | **Age 17** |  | **Age 24** |  |
| --- | --- | --- | --- | --- | --- |
|  |  | **RRR** | **95% CI** | **RRR** | **95% CI** |
| **Low parental education** | **Ref: 0 ACEs** | 1 |  | 1 |  |
|  | **1 ACE** | 0.74 | 0.30,1.84 | 0.80 | 0.38,1.69 |
|  | **2 to 3 ACEs** | 2.17 | 1.06,4.42 | 2.01 | 1.07,3.79 |
|  | **4 or more ACEs** | 4.19 | 1.88,9.34 | 3.11 | 1.49,6.51 |
| **High parental education** | **Ref: 0 ACEs** | 1 |  | 1 |  |
|  | **1 ACE** | 1.16 | 0.36,3.80 | 0.34 | 0.10,1.13 |
|  | **2 to 3 ACEs** | 1.17 | 0.37,3.73 | 1.01 | 0.43,2.40 |
|  | **4 or more ACEs** | 2.27 | 0.51,10.11 | 2.56 | 0.88,7.44 |
| **P-value for interaction** |  | 0.5576 |  | 0.7528 |  |

Note: Models are adjusted for gender, ethnicity, social class, financial difficulties and maternal age. ACE=adverse childhood experiences, RRR=relative risk ratio, CI=confidence interval.
